# Molecular detection and genetic characterisation of a large flood-borne outbreak of human leptospirosis in Jakarta, Indonesia: a retrospective analysis of surveillance data

**DOI:** 10.1101/2025.10.14.25338029

**Authors:** Erni Juwita Nelwan, Yunita Windi Anggraini, Sabighoh Zanjabila, Budi Setiawan, Suhartiningsih, Farida Dwi Handayani, Jeny, Linda Erlina, Fadilah, J. Kevin Baird, Raph L. Hamers, Suwarti

**Affiliations:** Division of Tropical and Infectious Diseases, Department of Internal Medicine, Cipto Mangunkusumo National Hospital, Jakarta, Indonesia; Infectious Disease and Immunology Research Center, Indonesian Medical Education and Research Institute, Universitas Indonesia, Jakarta, Indonesia; Oxford University Clinical Research Unit Indonesia, Faculty of Medicine, Universitas Indonesia, Jakarta, Indonesia; Jakarta Health Office, Jakarta, Indonesia; Regional Health Laboratory, Jakarta, Indonesia; Eijkman Research Center for Molecular Biology-Indonesian National Research and Innovation Agency, Jakarta, Indonesia; Bioinformatics Core Facilities, Indonesian Medical Education and Research Institute (IMERI), Universitas Indonesia, Jakarta, Indonesia; Department of Medical Chemistry, Faculty of Medicine, Universitas Indonesia, Jakarta, Indonesia; Centre for Tropical Medicine and Global Health, Nuffield Department of Medicine, University of Oxford, Oxford, United Kingdom

## Abstract

Recurring outbreaks of leptospirosis in flood-prone areas caused by heavy rainfall pose a major public health concern, particularly in megacities such as Indonesia’s capital city, Jakarta. From December 2019 through February 2020, Jakarta experienced a large leptospirosis outbreak due to extensive flooding following extreme monsoonal rainfall. We conducted a comprehensive retrospective analysis of the outbreak based on complete surveillance data from all five districts and 42 of 44 subdistricts in Jakarta. A total of 282 cases (97 suspected, 153 probable, and 32 confirmed) were reported in West (n=162), South (n=64), East (n=30), North (n=14) and Central (n=12) Jakarta. Cases were predominantly adult males exposed to floodwaters. Of 241 cases tested, 164 (68.0%) had a positive IgM-based rapid diagnostic test (RDT). Of 118 cases tested with TaqMan RT-PCR *lipL32*, 32 (27.1%) were positive; RT-PCR detected an additional 5 cases who were RDT-negative (case detection increased by 4.2% [5/118]), all of whom had fever <7 days. Of 95 cases tested with both assays, the combined detection rate was 74.7% (71/95). We sequenced 42 archived blood samples using Multi Locus Sequence Typing (MLST) and identified *Leptospira interrogans* and *L. borgpeterseni* as the predominant species. The findings emphasise the importance of rapid and early laboratory-based diagnosis during leptospirosis outbreaks in flood-prone urban areas, to better target public health interventions. Climate-resilient urban planning is critical for vulnerable megacities in low-resource settings, where complex environmental and infrastructural challenges are compounded by the effects of a changing climate.

**Author Summary:** Recurring leptospirosis outbreaks in flood-prone areas caused by heavy rainfall pose a major public health concern in vulnerable urban areas in low- and middle-income countries. This study investigated a large leptospirosis outbreak following extreme seasonal rainfall from December 2019 through February 2020 in Jakarta, Indonesia’s capital city. A total of 282 cases were identified, mostly adult males exposed to floodwaters. Combining point-of-care rapid diagnostic tests with RT-PCR laboratory-based tests improved early case detection. The *Leptospira* species identified were *L. interrogans* and *L. borgpetersenii*. This comprehensive analysis highlights the urgent need for improved diagnostic tests and surveillance systems, and enhanced disease control strategies. Climate-resilient urban planning is critical for vulnerable megacities where complex environmental and infrastructural challenges are compounded by the effects of a changing climate, including increased rainfall intensity.

## Introduction

Leptospirosis, a neglected zoonotic disease commonly found in tropical and sub-tropical regions, including Indonesia, is caused by spirochetes of the genus *Leptospira* spp., transmitted through water or soil contaminated by urine from infected livestock or rodents that serve as reservoirs [1, 2]. Jakarta, Indonesia’s capital city of 11 million populace and one the world’s megacities, located on Java Island, frequently experiences flooding during prolonged heavy seasonal rainfall [3]. These seasonal floods, coupled with high *Leptospira* spp. carriage in sewer rats is a major driver of leptospirosis outbreaks [4]. Contributing environmental and infrastructural challenges include inadequate sanitation and drainage systems, low elevation worsened by rapid land subsidence, high population density and insufficient public health infrastructure. These are amplified by the emerging effects of climate change through rising sea levels, increased rainfall intensity and more frequent extreme weather events [5]. The end of December 2019 saw the most significant rainfall in the last 24 years (reaching a high of 377 millimetres per day), causing recurrent floods up to the end of February 2020 [6]. On January 2, 2020, authorities declared an official public health alert following reports of human leptospirosis cases [7].

The Ministry of Health of Indonesia has implemented a surveillance system for the early detection of leptospirosis outbreaks, mainly relying on clinical case reports because of limited laboratory diagnostic capacity [8]. However, the diverse and non-specific clinical manifestations of leptospirosis lead to frequent misdiagnosis and underreporting [9]. Between 2019 and 2023, the annual number of leptospirosis cases ranged from 736 to 2554 nationwide and from 15 to 209 in Jakarta [10].

Although several surveillance studies have been conducted in Indonesia [11–13], there is a lack of laboratory-based case detection and confirmation during outbreaks. Reference diagnostic methods, such as the Microscopic Agglutination Test (MAT) and bacterial cultures, are expensive, require specialised expertise [14, 15] and have limited availability in Indonesia [11]. Molecular diagnostic, Real-time PCR (RT-PCR) targeting the *lipL32* gene has been used in outbreak settings as a practical diagnostic tool for early detection and increased case identification [16, 17], while Multi-locus sequence typing (MLST) is a useful method to identify pathogenic *Leptospira* species in clinical samples [16, 17].

This study aimed to 1) describe the leptospirosis outbreak in Jakarta based on the complete government surveillance data from December 2019 through February 2020; 2) evaluate the performance of RDT and TaqMan RT-PCR *lipL32* as diagnostic tools during the outbreak; and 3) characterise circulating pathogenic *Leptospira* species using MLST.

## Methods

### Study design and study population

We performed a retrospective analysis of the complete surveillance data reported to the Jakarta Health Office from 21 December 2019 through 28 February 2020. Data were obtained from 24 primary healthcare centres, 16 public hospitals, and 15 private hospitals covering the five districts (West, South, East, North, and Central) and 42 of 44 flood-affected subdistricts. The two subdistricts not included, located in Kepulauan Seribu islands, did not report any leptospirosis cases.

The surveillance system recorded suspected, probable, and confirmed leptospirosis cases in accordance with case definitions of the Indonesian Ministry of Health [8]. Suspected case was defined as acute fever (≥ 37.5 °C) with or without headaches accompanied by muscle pain, malaise, or conjunctival suffusion, and a history of exposure to risk factors (in this case, flood or water puddle) within the past two weeks. Probable case was defined as a suspected case with: i) two or more of the following clinical symptoms: calf pain, icterus, oliguria, haemorrhage, dyspnoea, arrhythmia, cough with or without haemoptysis, and skin rash; or ii) a positive RDT; or iii) at least three of the following laboratory results: thrombocytopenia (<100.000 cells/mm^3^), neutrophilia (>80%), elevated total serum bilirubin, and proteinuria. Confirmed case was defined as a suspected case or probable case, with at least one of the following additional diagnostic test results: i) positive *Leptospira* culture from a clinical specimen; ii) positive PCR; iii) seroconversion from negative to positive or 4-fold titre rise from baseline to convalescent timepoint with MAT.

### Ethics statement

The study was approved by the Research Ethical Committee of the Faculty of Medicine Universitas Indonesia (19-05-0608) and the Oxford Tropical Research Ethics Committee (33-19). This study is reported as per the STrengthening the Reporting of OBservational studies in Epidemiology (STROBE) guidelines [18].

### Data collection

A case record form captured case definition data, date of symptom onset, age, sex, flood exposure, fever onset, clinical symptoms, complete blood cell count and urinalysis. Leptospirosis diagnostic tests were conducted as part of surveillance with IgM serological Rapid Diagnostic Test (RDT) (Pakar Biomedika, Indonesia) in each health care facility and TaqMan RT-PCR targeting the *lipL32* gene in the centralised Jakarta Regional Health Laboratory [8]. Daily precipitation data were obtained from the open data on the meteologix website [19], flood data from Indonesia’s National Disaster Management Agency (BNPB) via Satudata [20], and district-level population data from BPS Statistics of Jakarta Province [21].

### Laboratory analysis

We conducted MLST and phylogenetic analysis of the residual whole-blood samples obtained for surveillance purposes and archived at the Jakarta Regional Health Laboratory. The total genomic DNA was extracted from 200µL whole blood using DNAeasy Blood and Tissue (Qiagen, Germany). MLST of the extracted DNA was performed through conventional PCR targeting *adk*, *icdA, lipL32, lipL41, rrs*, and *secY* genes in accordance with MLST scheme 3 [22]. All primers used were listed in Table S1. PCR amplification was conducted for 40 cycles using GoTaq® Green Master Mix in a 40 μL volume on a SimpliAmp Thermal Cycler (Applied Biosystem). Amplicons were visualised on a 2% agarose gel stained with SYBR Safe DNA gel stain (Invitrogen) and subsequently sequenced by First Base Laboratories (Selangor, Malaysia). All sequenced amplified molecular markers were concatenated. MLST allele and sequence types (STs) assignments were determined for samples with ≥4 positive loci, assigning them to the closest matching STs. Phylogenetic analysis of all sequences was performed using the Maximum-Likelihood tree construction method with 1000 bootstrap replicates and the General Time Reversible Gamma distribution Invariable sites (GTR+G+I) model, including reference sequences from the PubMLST database.

### Data Analysis

Data analysis was conducted in R version 4.4.3 (R Core Team, Vienna, Austria). Data were presented as frequencies (%) and medians (IQR). District and subdistrict-level leptospirosis incidence rates were expressed per 100,000 person-years, calculated as the number of cases during the 3-month observation period divided by population size (December 2019 to February 2020). Geographic case mapping was plotted using QGIS version 3.32 with a basemap by the Geospatial Information Agency of Indonesia (https://geoservices.big.go.id). Test positivity and diagnostic yield of diagnostic tests were expressed as n/N (%). Gene alignment, concatenation, and multi-locus sequence typing (MLST) analysis were performed using a custom homebrew pipeline via command-line tools and analysed based on MLST scheme 3 from the PubMLST database (https://pubmlst.org/leptospira/). The phylogenetic tree was constructed in R version 4.4.3 (R Core Team, Vienna, Austria).

## Results

### Geospatial and temporal description of the outbreak

**Fig 1** shows the timeline of the outbreak. The first probable case was reported on December 21, 2019. The outbreak peaked from January 10–14, 2020, about two weeks after the severe flooding. A second flooding event began on January 18, 2020, lasting nearly a week, triggering a secondary outbreak. The outbreak gradually declined by the end of January, with the final cases reported on February 28, 2020.

**Fig 1.**
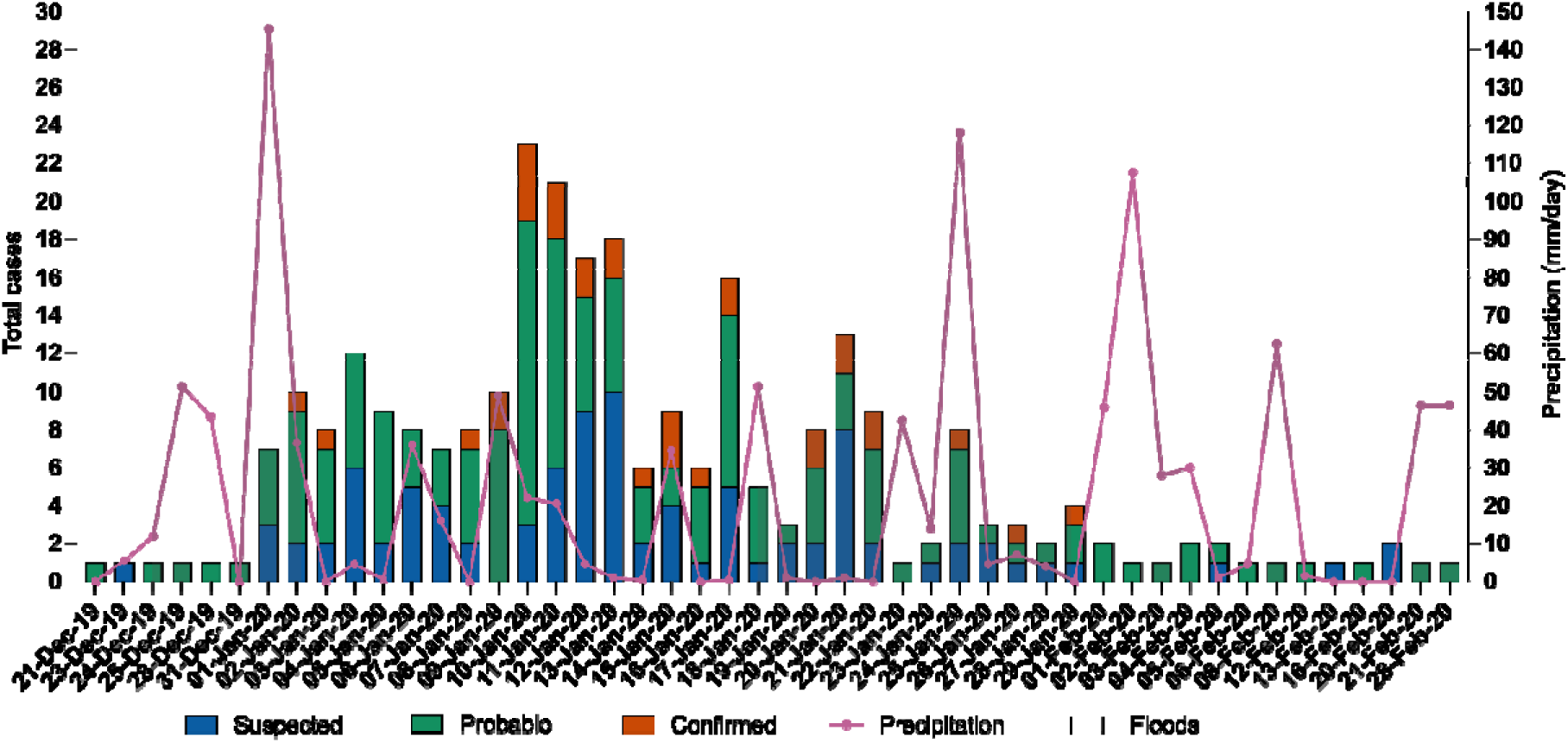
The epidemic curve of the leptospirosis outbreak in Jakarta with daily precipitation data during the rainy season from December 2019 to February 2020. Bars represent suspected (blue), probable (green), and confirmed (red) cases recorded by the surveillance. The left y-axis expresses the total daily cases of leptospirosis, and the right y-axis expresses the daily total precipitation (mm/day). The upper horizontal bar indicates the flood period.

All five districts of Jakarta were affected by floods (35 of 42 [83.3%] subdistricts) and reported leptospirosis cases (33 of 42 [78.6%] subdistricts). The overall incidence rate in Jakarta was 10.7 per 100,000 person-years. West Jakarta had the highest leptospirosis incidence (n=162; 26.6 per 100,000 person-years), with 6 of 8 subdistricts reporting cases (top-3: Cengkareng n=72, Kalideres n=23, Kebon Jeruk n=22); followed by South Jakarta (n=64; 11.5 per 100,000 person-years), across all 10 subdistricts (top-3: Tebet n=16, Cilandak, and Kebayoran Baru n=10 each); Central Jakarta (n=12; 4.5 per 100,000 person-years), with 4 of 8 subdistricts reporting cases (top-3: Sawah Besar, Tanah Abang n=4 each; and Kemayoran n=2); East Jakarta (n=30; 4.0 per 100,000 person-years), with 7 of 10 subdistricts reporting cases (top-3: Kramat Jati n=10, Jatinegara n= 8, and Makasar n=5); and North Jakarta (n=14; 3.1 per 100,000 person-years), with 5 of 6 subdistricts reporting cases (top-3: Penjaringan, Tanjung Priok (a non-flooded subdistrict) each n=4; and Cilincing n=3). Notably, some non-flooded subdistricts, such as Tanjung Priok in North Jakarta and Tambora in West Jakarta, reported cases, while some flooded subdistricts, such as Kelapa Gading in North Jakarta and Cakung, Ciracas, and Cipayung in East Jakarta, did not report any cases (**Fig 2, Table S2**).

**Fig 2.**
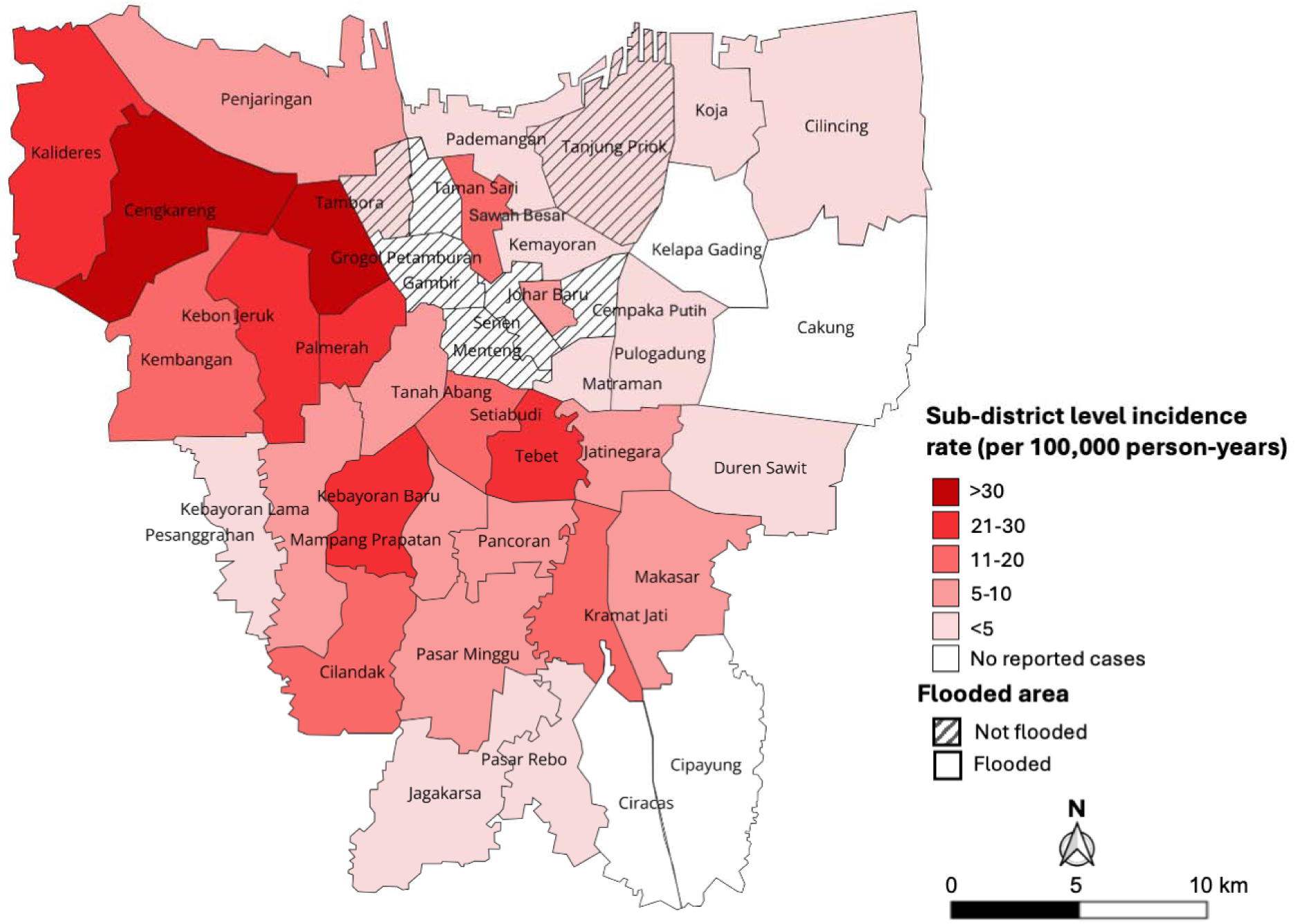
Incidence rate of leptospirosis (per 100,000 person-years) and spatial distribution of flood-affected sub-district during the heavy flood period (December 2019-February 2020) in Jakarta, Indonesia. The gradient from white to red represents the incidence rate, while not-flooded subdistricts are shaded. Maps were created in QGIS version 3.32 using basemaps from the Geospatial Information Agency of Indonesia (https://geoservices.big.go.id).

### Patient characteristics

Of the 285 cases, three cases had incomplete clinical and laboratory data; therefore, 282 cases were included in the analysis, as illustrated in the **Fig 3**. The cases were classified as suspected (97, 34.4%), probable (153, 54.3%) or confirmed (32,11.3%). Cases were predominantly male (n=203, 72.0%), and aged 18 to 60 years (n=212, 75.2%), and reported flood exposure (n=228, 80.9%) within the preceding two weeks (**Table 1**).

**Fig 3.**
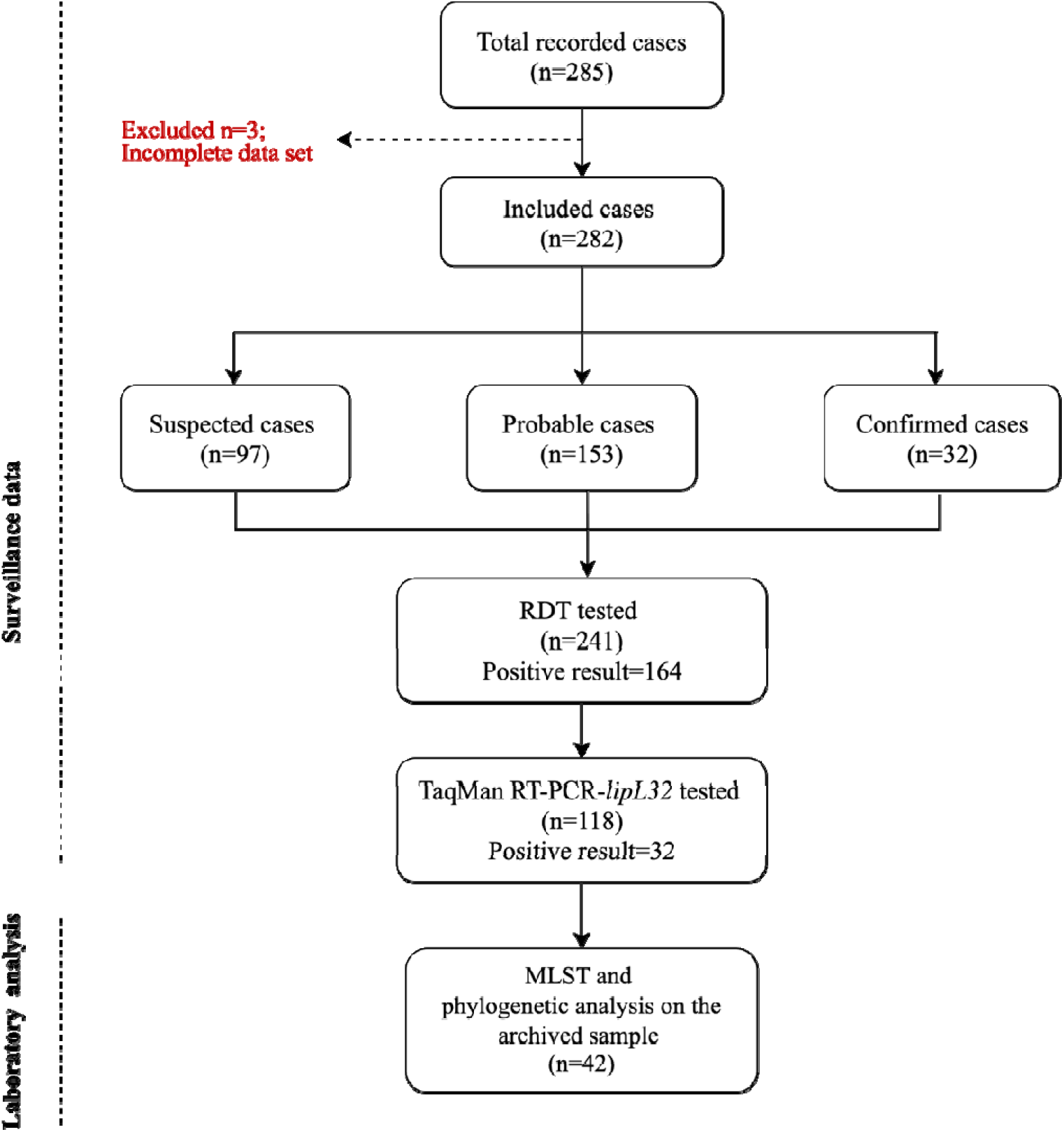
Study flow diagram. Abbreviations: MLST, multi-locus sequence typing; PCR, polymerase chain reaction; RDT, rapid diagnostic test; RT-PCR, real-time polymerase chain reaction.

**Table 1.** Patient characteristics and laboratory test results.

| Characteristics | Total (%) | Cases (%) |  |  |
| --- | --- | --- | --- | --- |
|  |  | Suspected | Probable | Confirmed |
| Number of cases | 282 (100) | 97 (34.4) | 153 (54.3) | 32 (11.3) |
| Age - Median (IQR) years | 38 (24–51) | 37 (24–49) | 39 (23–52) | 43 (33–55) |
| **Age group** |  |  |  |  |
| Children (1-12 years) | 23 (8.2) | 7 (7.2) | 14 (9.2) | 2 (6.3) |
| Adolescents (13-17 years) | 14 (5.0) | 5 (5.2) | 8 (5.2) | 1 (3.1) |
| Adults (18-60 years) | 212 (75.2) | 75 (77.3) | 112 (73.2) | 25 (78.1) |
| Elderly (>60 years) | 33 (11.7) | 10 (10.3) | 19 (12.4) | 4 (12.5) |
| **Sex** |  |  |  |  |
| Male | 203 (72.0) | 31 (32.0) | 43 (28.1) | 5 (15.6) |
| Female | 79 (28.0) | 66 (68.0) | 110 (71.9) | 27 (84.4) |
| **Occupation** |  |  |  |  |
| Employee/self-employee | 92 (32.6) | 31 (32.0) | 51 (33.3) | 10 (31.3) |
| Unemployed | 52 (18.4) | 17 (17.5) | 28 (18.3) | 7 (21.9) |
| Labourer | 50 (17.7) | 14 (14.4) | 28 (18.3) | 8 (25.0) |
| Housewife | 27 (9.6) | 8 (8.2) | 16 (10.5) | 3 (9.4) |
| Student | 21 (7.4) | 9 (9.3) | 12 (7.8) | 0 (0.0) |
| Unspecified | 40 (14.2) | 18 (18.6) | 18 (11.8) | 4 (12.5) |
| **District** |  |  |  |  |
| West Jakarta | 162 (57.4) | 60 (61.9) | 92 (60.1) | 10 (31.3) |
| South Jakarta | 64 (22.7) | 24 (24.7) | 33 (21.6) | 7 (21.9) |
| East Jakarta | 29 (10.3) | 8 (8.2) | 18 (11.8) | 3 (9.4) |
| North Jakarta | 14 (5.0) | 1 (1.0) | 6 (3.9) | 7 (21.9) |
| Central Jakarta | 13 (4.6) | 4 (4.1) | 4 (2.6) | 5 (15.6) |
| **Fever onset** |  |  |  |  |
| Overall | 211 (74.8) | 68 (70.1) | 118 (77.1) | 25 (78.1) |
| < 7 days | 125 (44.3) | 40 (41.2) | 67 (43.8) | 18 (56.3) |
| ≥ 7 days | 86 (30.5) | 28 (28.9) | 51 (33.3) | 7 (21.9) |
| Unspecified | 71 (25.2) | 29 (29.9) | 35 (22.9) | 7 (21.9) |
| **Risk factor** |  |  |  |  |
| Exposed to flood in the past 2 weeks | 228 (80.9) | 73 (75.3) | 126 (82.4) | 29 (90.6) |
| Unexposed to flood in the past 2 weeks | 29 (10.3) | 11 (11.3) | 17 (11.1) | 1 (3.1) |
| Unspecified | 25 (8.9) | 13 (13.4) | 10 (6.5) | 2 (6.3) |
| **Leptospirosis diagnostic test** |  |  |  |  |
| Rapid Diagnostic Test (RDT) | 241 (85.5) | 65 (67.0) | 147 (96.1) | 29 (90.6) |
| Positive | 164 (68.0) | 0(0.0) | 140 (95.2) | 24 (75.0) |
| Negative | 77 (32.0) | 65 (100) | 7 (4.6) | 5 (15.6) |
| Not done | 41 (14.5) | 32 (33.0) | 6 (3.9) | 3 (9.4) |
| TaqMan RT-PCR *lipL32* | 118 (41.8) | 39 (40.2) | 47 (30.7) | 32 (100) |
| Positive | 32 (27.1) | 0 (0.0) | 0 (0.0) | 32 (100.0) |
| Negative | 86 (72.8) | 39 (100.0) | 47 (100.0) | 0 (0.0) |
| Not done | 163 (57.8) | 58 (59.8) | 105 (68.6) | 0 (0.0) |
| Either RDT and/or TaqMan RT-PCR<br>*lipL32* | 95 (33.7) | 20 (20.6) | 46 (30.1) | 29 (90.6) |
| Positive | 71 (74.7) | 0 (0.0) | 42 (91.3) | 29 (100) |
| Negative | 24 (25.3) | 20 (100.6) | 4 (8.7) | 0 (0.0) |
| Unrecorded in any tests | 18 (6.4) | 13 (13.4) | 5 (3.3) | 0 (0.0) |

Data are n (%) unless specified otherwise
Abbreviations: IQR, interquartile range; RT-PCR, real-time polymerase chain reaction.

**Table 1** summarises the patient characteristics. The most common characteristics were fever, myalgia, malaise, neutrophilia, calf pain, and conjunctival suffusion (**Fig 4**).

**Fig 4.**
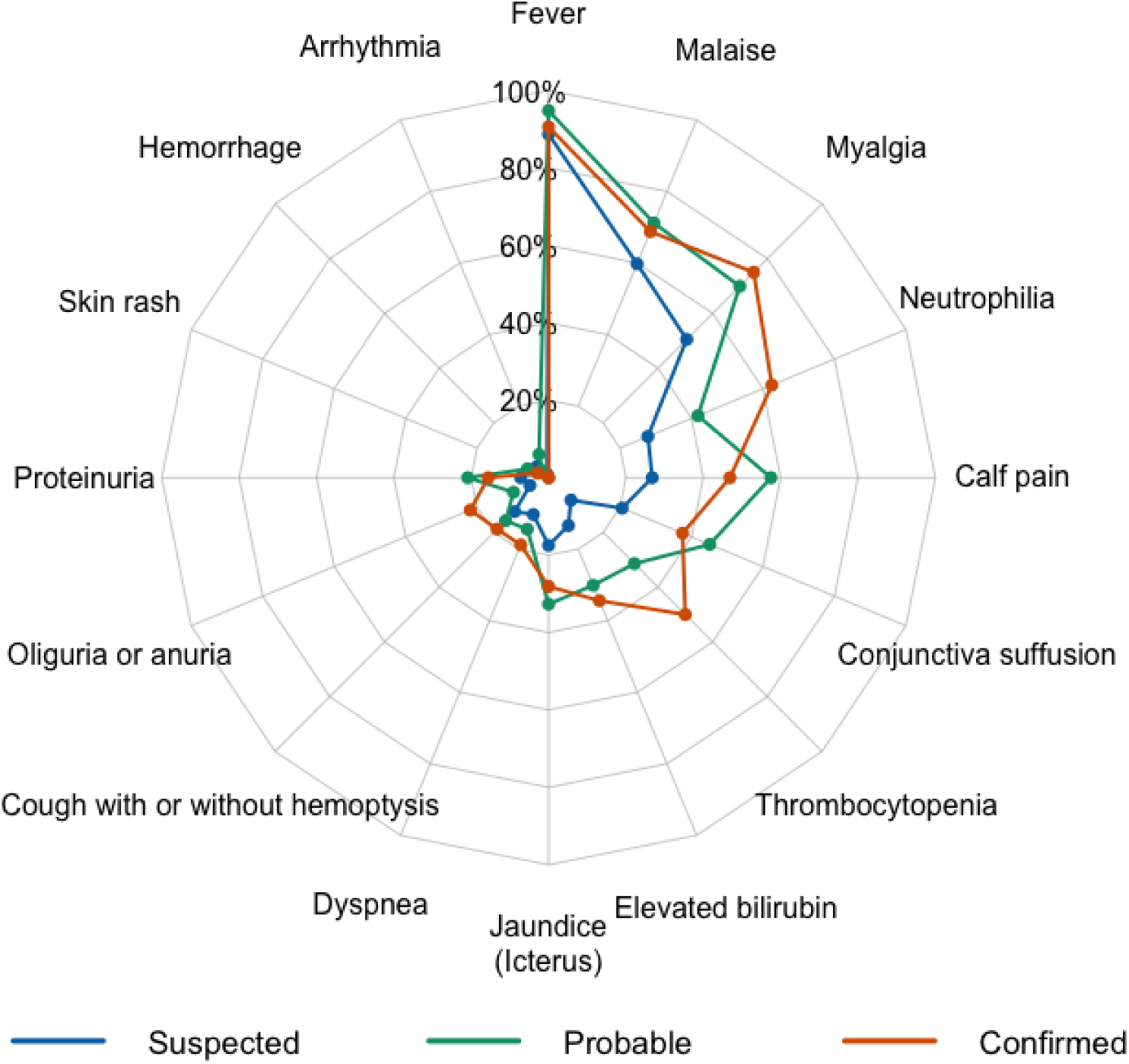
**Spider chart showing clinical characteristics of the 282 leptospirosis cases**

### Diagnostic test performance

Of 241 patients tested with RDT, 68.0% (164/241) were RDT-positive. Of 118 tested with TaqMan RT-PCR *lipL32*, 27.1% (32/118) were positive. Among the 282 individuals, 95 (33.7%) were tested using both RDT and RT-PCR targeting *lipL32*, while 18 (6.4%) were not tested by either method (**Table 1**, **Fig. 3**). RT-PCR detected an additional 5 cases who were RDT-negative (diagnostic yield of 5/118 [4.2%]), all of whom had fever <7 days (**Fig 5**), yielding a combined test positivity of 74.7% (71/95) (**Table 1**).

**Fig 5.**
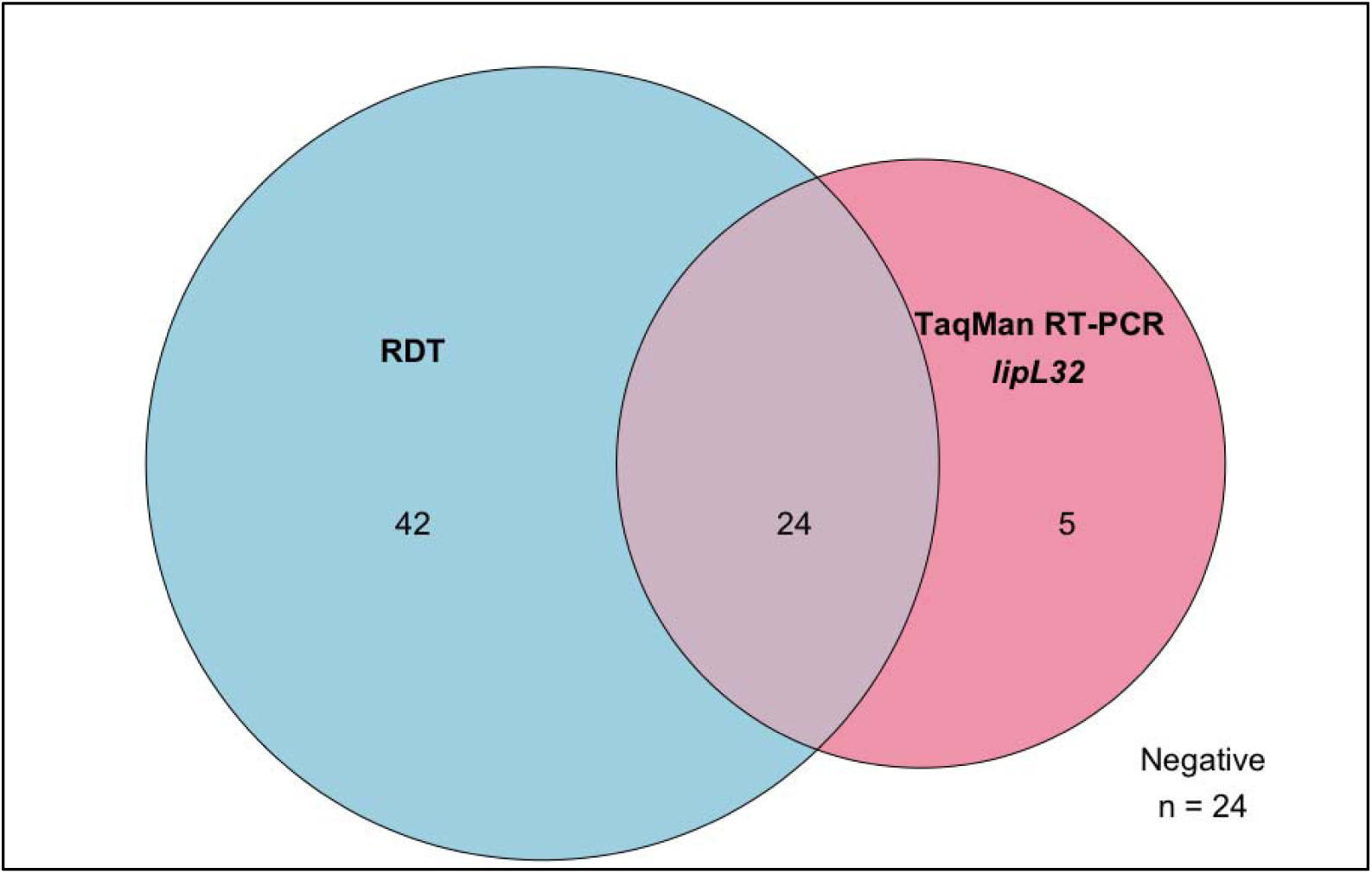
Venn diagram illustrating the overlap for positive tests among patients tested with both RDT and TaqMan RT-PCR *lipL32* (n=95). Of 95 patients who received both tests, 42 patients tested positive by RDT only, 5 by RT-PCR only, and 24 were positive by both methods.

### *Leptospira* spp. identification and phylogenetic evaluation

PCR analysis on DNA from 42 archived whole-blood samples identified the *secY* gene in 15 samples (35.7%), *lipL32* in 13 (31%), *rrs* in 7 (16.7%), *adk* in 6 (14.3%), *lipL41* in 6 (14.3%), and *icdA* in 3 (7.1%). Overall, 17 samples (40.5%) were confirmed positive for leptospirosis through the detection of at least one of these MLST genes (**S3 Table**). However, MLST scheme 3 was only able to assign the allele for 5 samples (01, 04, 14, 55, and 71) (**Table 2**). Among those five, two were identified as sequence types (STs) of *L. borgpetersenii* (STs 193 or 194) and sample 71 forming a distinct branch within the *L. interrogans* cluster (**Table 2**, **Fig 6**).

**Fig 6.**
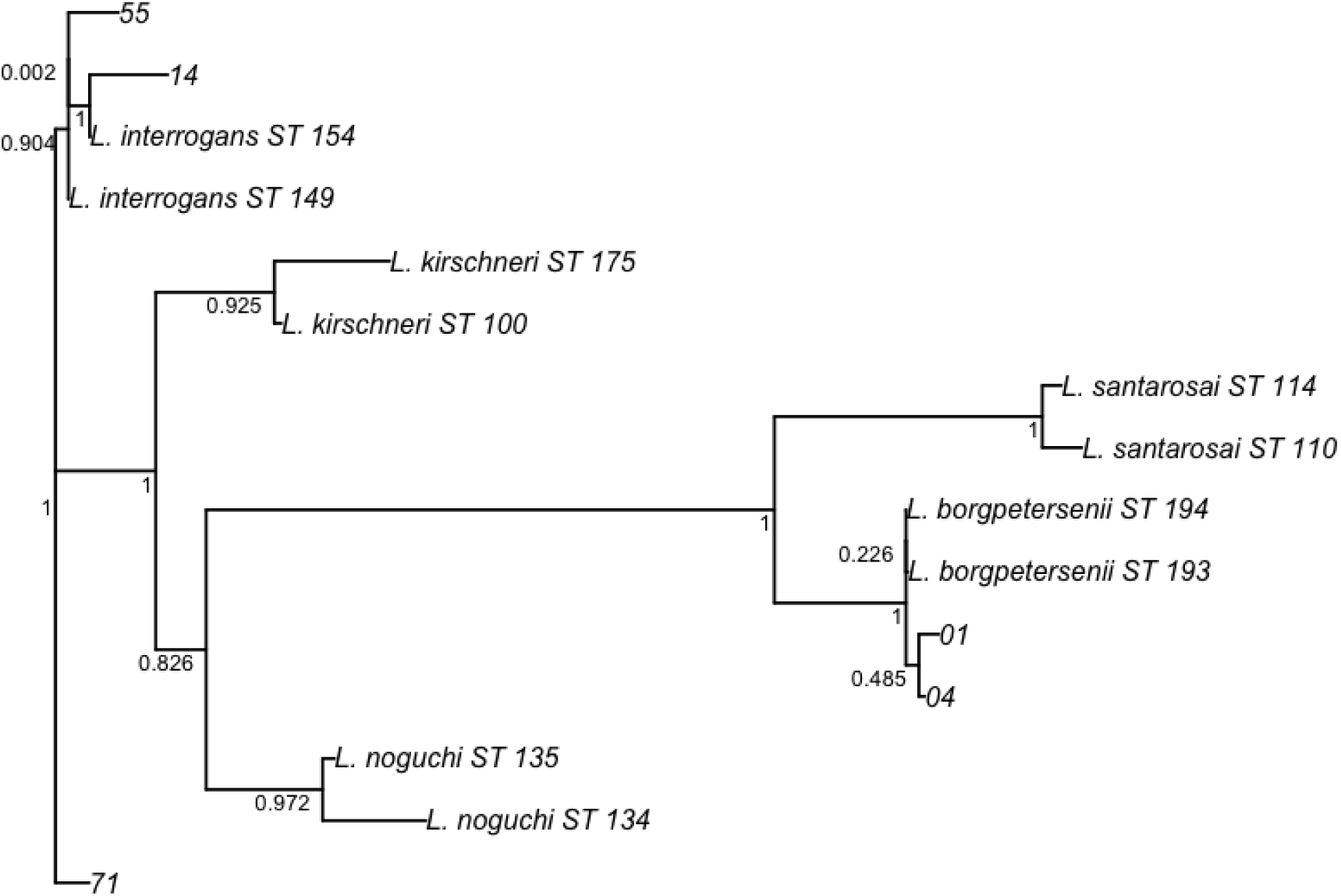
Maximum Likelihood tree of *Leptospira* using concatenated sequences from MLST scheme 3 loci gene sequences alignment, constructed under GTR+G+I substitution model. Clinical samples (01, 04, 14, 55, and 71) are shown in relation to commonly reported reference pathogenic *Leptospira* sequence types (STs) from the PubMLST database. Bootstrap values (×1000) are displayed at the branch nodes to demonstrate the reliability of phylogenetic relationships. Corresponding sequence type isolate IDs and allele information from the PubMLST database are provided in **S4 Table**.

**Table 2.** The result of MLST, allele assignation, presumptive STs and serovar of *Leptospira* in clinical samples.

| Sample ID | Alleles assignation |  |  |  |  |  | Presumptive STs*<br>(MLST scheme 3) | Species | Presumptive serovar** |
| --- | --- | --- | --- | --- | --- | --- | --- | --- | --- |
|  | *adk* | *icdA* | *lipL32* | *lipL41* | *rrs* | *secY* |  |  |  |
| 01 | - | - | 10 | 15 | 30 | 49 | 193 or 194 | *L. borgpetersenii* | NA |
| 04 | - | - | 10 | 15 | 30 | 49 | 193 or 194 | *L. borgpetersenii* | NA |
| 14 | 5 | 2 | 2 | 50 | - | 13 | 154 | *L. interrogans* | NA |
| 55 | 5 | 1 | 2 | 3 | - | 5 | 149 | *L. interrogans* | Bataviae/Canicola |
| 71 | 5 | 1 | 2 | 3 | - | 5 | 149 | *L. interrogans* | Bataviae/Canicola |

- Indicates no amplification
\* Samples with an allelic profile were assigned to the nearest match ( $\geq 4$ matching loci) of sequence type (ST) in the *Leptospira* PubMLST database
(https://pubmlst.org/leptospira/)
\* \* The presumptive serovars were determined based on sequence type (ST) matching in the *Leptospira* PubMLST database
Abbreviations: ID, identity; MLST, multi-locus sequence typing; NA, not applicable; ST, sequence type.

## Discussion

To our knowledge, this is the first comprehensive molecular analysis of a large leptospirosis outbreak in Indonesia, identifying *L. interrogans* and *L. borgpetersenii* as the main circulating *Leptospira* species. Most (68%) cases were RDT-positive, and the addition of TaqMan RT-PCR *lipL32* increased case detection by 4.2%.

Flood-related leptospirosis outbreaks are common in tropical and sub-tropical regions, often driven by intense [23] or prolonged rainfall [24, 25], facilitating waterborne transmission, compounded by environmental and infrastructural challenges and the effects of a changing climate [5].

Our study observed that leptospirosis cases peaked 10–14 days after heavy rainfall, with outbreaks occurring within 2–4 weeks post-flooding [26, 27]. The predominance of middle-aged males is consistent with studies elsewhere, reflecting a higher risk of exposure to contaminated floodwaters [25, 26, 28]. Our findings underscore the importance of increased clinical awareness in flood-prone areas during the rainy season and access to rapid diagnostic tests for early outbreak detection, and prompt diagnosis and treatment [29].

*Leptospira* spp. culture is unreliable in outbreak settings due to slow bacterial growth, while MAT is limited to a few referral laboratories, further delaying diagnosis and treatment. Serology-based tests, like ELISA and RDT, require the accumulation of detectable amounts of anti-*Leptospira* antibodies, typically present in late acute to convalescent samples (10–14 days post-infection) [30]. Although currently available RDTs have a wide variation in reported sensitivity (68%–93%), it remains an essential tool for accessible, point-of-care diagnosis during outbreaks, particularly in settings with limited laboratory capacity [16, 31]. Add-on RT-PCR targeting the *lipL32* gene has been reported to improve case detection and confirmation by 3–6% [32, 33], particularly in the first 3–8 days after symptom onset, which was consistent with our findings [34]. Variation in diagnostic yield across studies may reflect variability in patient populations, timing of sampling, sample quality, and assay performance, highlighting the need to further improve the implementation of current RT-PCR protocols.

This study identified *L. interrogans* and *L. borgpetersenii* as the dominant pathogenic species, which are also the two most abundant species detected in rodent populations and human disease in Asia [35, 36]. Most studies in Southeast Asia have focused on human infection in rural settings, like rice cultivation and other humid habitats. Our study highlights infection risks in flood-prone, densely populated urban environments, where *Leptospira*-carrying rats live in the sewage system [35]. In addition, *L. interrogans,* with serovars Bataviae and Canicola, have been found to be predominant in domestic animals [37], indicating that stray dogs and cats may serve as an additional reservoir during urban floods. We did not detect any other species, for example, *L. kirschneri*, *L. wolffii*, *L. santarosai*, and *L. weilii,* which have been previously reported in outbreaks in Southeast Asia [35, 38].

There are several limitations to this study. First, we relied on routinely collected surveillance data. This meant that some demographic, clinical, and laboratory data were incomplete and that we could not assess patient outcomes. Second, surveillance priorities shifted to the COVID-19 pandemic in March 2020, leaving the possibility that later cases were unrecorded. Third, we were not able to conduct leptospirosis reference testing, by culture and MAT, on the samples due to limited resources. Lastly, because we only genetically characterised a subset of the pathogens, we may have missed additional *Leptospira* species.

In conclusion, this analysis illustrates the risks of large leptospirosis outbreaks in vulnerable megacities where the complex interaction of infectious diseases, environment, infrastructure, and climate change presents formidable challenges. There is an urgent need for improved diagnostic tests and surveillance systems, enhanced disease control strategies and climate-resilient urban planning.

## Supporting Information

Table S1. List of primers used in this study

Table S2. Distribution of the leptospirosis cases across the districts and subdistricts of Jakarta (December 2019 to February 2020).

Table S3. PCR-positive samples for *Leptospira* in six MLST genes.

Table S4. Reference isolates from the *Leptospira* PubMLST database used in phylogenetic tree construction, including sequence types (STs) and allele profiles based on MLST scheme 3.

## Data Availability

All data produced in the present work are contained in the manuscript

## Acknowledgements

The authors express their gratitude to the Jakarta Health Office, the Ministry of Health of Indonesia, and the healthcare professionals who contributed to the data collection. The authors were also grateful to Kartika Saraswati, PhD and Made Ananda Krisna,PhD, for their valuable input during the development of this paper.

## Author contributions

**Conceptualisation:** Suwarti, Erni Juwita Nelwan, Raph L. Hamers

**Data curation:** Budi Setiawan, Suhartiningsih

**Formal analysis:** Yunita Windi Anggraini, Sabighoh Zanjabila, Linda Erlina, Fadilah

**Funding acquisition:** J. Kevin Baird, Raph L. Hamers, Erni Juwita Nelwan, Suwarti

**Investigation:** Yunita Windi Anggraini, Sabighoh Zanjabila, Jeny, Suwarti

**Methodology:** Yunita Windi Anggraini, Sabighoh Zanjabila, Jeny, Suwarti, Farida Dwi Handayani

**Project administration:** Sabighoh Zanjabila

**Supervision:** Suwarti, Erni Juwita Nelwan, Raph L. Hamers

**Visualisation:** Yunita Windi Anggraini, Sabighoh Zanjabila, Suwarti, Raph L. Hamers, Linda Erlina, Fadilah

**Writing – original draft:** Yunita Windi Anggraini

**Writing – review& editing:** Suwarti, Raph L. Hamers, Erni Juwita Nelwan, J. Kevin Baird, Farida Dwi Handayani

