## Supplementary file for "Molecular detection and genetic characterisation of a large flood-borne outbreak of human leptospirosis in Jakarta, Indonesia: a retrospective analysis of surveillance data"

**Table S1.** List of primers used in this study

| Genes | Primers | PCR product size (bp) |
|---|---|---|
| *adk* | Forward: 5'-GGG CTG GAA AAG GTA CAC AA-3'<br>Reverse: 5'-ACG CAA GCT CCT TTT GAA TC-3' | 531 |
| *icdA* | Forward: 5'-GGG ACG AGA TGA CCA GGA T-3'<br>Reverse: 5'-TTT TTT GAG ATC CGC AGC TTT-3' | 674 |
| *lipL32* | Forward: 5'-ATC TCC GTT GCA CTC TTT GC-3'<br>Reverse: 5'-ACC ATC ATC ATC ATC GTC CA-3' | 474 |
| *lipL41* | Forward: 5'-TAG GAA ATT GCG CAG CTA CA-3'<br>Reverse: 5'-GCA TCG AGA GGA ATT AAC ATC A-3' | 520 |
| *rrs* | Forward: 5'-GGC GGC GCG TCT TAA ACA TG-3'<br>Reverse: 5'-TTC CCC CCA TTG AGC AAG ATT-3' | 331 |
| *secY* | Forward: 5'- ATG CCG ATC ATT TTT GCT TC-3'<br>Reverse: 5'- CCG TCC CTT AAT TTT AGA CTT CTT C-3' | 549 |

**Table S2.** Distribution of the leptospirosis cases across the districts and subdistricts of Jakarta (December 2019 to February 2020).

| District-level data | | | | Subdistrict-level data | | | | |
|---|---|---|---|---|---|---|---|---|
| **District** | **Population size*** | **Number of cases** | **Incidence rate** (per 100,000 person-years)** | **Subdistrict** | **Flooded** | **Population size*** | **Number of cases** | **Incidence rate** (per 100,000 person-years)** |
| West Jakarta | 2,434,511 | 162 | 26.6 | Cengkareng | Yes | 551,682 | 72 | 52.2 |
| | | | | Kalideres | Yes | 430,575 | 23 | 21.4 |
| | | | | Kebon Jeruk | Yes | 341,938 | 22 | 25.7 |
| | | | | Grogol Petamburan | Yes | 228,669 | 21 | 36.7 |
| | | | | Palmerah | Yes | 217,310 | 13 | 23.9 |
| | | | | Kembangan | Yes | 288,768 | 8 | 11.1 |
| | | | | Tambora | No | 256,060 | 3 | 4.7 |
| | | | | Taman Sari | No | 119,509 | NA | NA |
| South Jakarta | 2,226,812 | 64 | 11.5 | Tebet | Yes | 221,191 | 16 | 28.9 |
| | | | | Cilandak | Yes | 201,591 | 10 | 19.8 |
| | | | | Kebayoran Baru | Yes | 139,592 | 10 | 28.7 |
| | | | | Pasar Minggu | Yes | 304,293 | 8 | 10.5 |
| | | | | Kebayoran Lama | Yes | 308,510 | 5 | 6.5 |
| | | | | Jagakarsa | Yes | 383,380 | 4 | 4.2 |
| | | | | Mampang Prapatan | Yes | 145,387 | 3 | 8.3 |
| | | | | Pancoran | Yes | 168,596 | 3 | 7.1 |
| | | | | Setiabudi | Yes | 107,392 | 3 | 11.2 |
| | | | | Pesanggrahan | Yes | 246,880 | 2 | 3.2 |
| Central Jakarta | 1,056,896 | 12 | 4.5 | Sawah Besar | Yes | 122,500 | 4 | 13.1 |
| | | | | Tanah Abang | Yes | 175,150 | 4 | 9.1 |
| | | | | Johar Baru | Yes | 133,713 | 2 | 6.0 |
| | | | | Kemayoran | Yes | 240,631 | 2 | 3.3 |
| | | | | Cempaka Putih | No | 94,031 | NA | NA |
| | | | | Gambir | No | 91,673 | NA | NA |
| | | | | Menteng | No | 80,319 | NA | NA |
| | | | | Senen | No | 118,879 | NA | NA |

| District-level data | | | | Subdistrict-level data | | | | |
|---|---|---|---|---|---|---|---|---|
| **District** | **Population size*** | **Number of cases** | **Incidence rate** (per 100,000 person-years)** | **Subdistrict** | **Flooded** | **Population size*** | **Number of cases** | **Incidence rate** (per 100,000 person-years)** |
| East Jakarta | 3,037,139 | 30 | 4.0 | Kramat Jati | Yes | 298,437 | 10 | 13.4 |
| | | | | Jatinegara | Yes | 301,717 | 8 | 10.6 |
| | | | | Makasar | Yes | 207,293 | 5 | 9.6 |
| | | | | Duren Sawit | Yes | 414,604 | 2 | 1.9 |
| | | | | Matraman | Yes | 172,180 | 2 | 4.6 |
| | | | | Pulo Gadung | Yes | 281,319 | 2 | 2.8 |
| | | | | Pasar Rebo | Yes | 220,583 | 1 | 1.8 |
| | | | | Cakung | Yes | 559,040 | NA | NA |
| | | | | Cipayung | Yes | 285,650 | NA | NA |
| | | | | Ciracas | Yes | 296,316 | NA | NA |
| North Jakarta | 1,778,981 | 14 | 3.1 | Penjaringan | Yes | 315,613 | 4 | 5.1 |
| | | | | Tanjung Priok | No | 401,806 | 4 | 4.0 |
| | | | | Cilincing | Yes | 428,316 | 3 | 2.8 |
| | | | | Pademangan | Yes | 162,843 | 2 | 4.9 |
| | | | | Koja | Yes | 331,616 | 1 | 1.2 |
| | | | | Kelapa Gading | Yes | 138,787 | NA | NA |

*The population size data was sourced from the BPS Statistics of Jakarta Province, 2020.

** Incidence rates were expressed per 100,000 person-years, calculated as the number of cases divided by the population and adjusted for the 3-month observation period (December 2019 to February 2020).

Abbreviation: NA, data were not reported

**Table S3.** PCR-positive samples for *Leptospira* in six MLST genes.

| **Sample ID** | ***adk*** | ***icdA*** | ***lipL32*** | ***lipL41*** | ***rrs*** | ***secY*** | **Positive genes amplified** |
|---|---|---|---|---|---|---|---|
| 01 | - | - | + | + | + | + | 4 |
| 03 | - | - | - | - | - | + | 1 |
| 04 | - | - | + | + | + | + | 4 |
| 06 | - | - | + | - | - | + | 2 |
| 08 | + | - | - | + | + | - | 3 |
| 09 | - | - | + | - | - | + | 2 |
| 12 | - | - | + | - | - | + | 2 |
| 14 | + | + | + | + | + | + | 6 |
| 17 | + | - | - | - | + | + | 3 |
| 28 | - | - | - | - | - | + | 1 |
| 45 | + | + | + | - | - | + | 4 |
| 55 | + | + | + | + | + | + | 6 |
| 62 | - | - | + | - | - | + | 2 |
| 65 | - | - | + | - | - | + | 2 |
| 66 | - | - | + | - | - | + | 2 |
| 69 | - | - | + | - | - | + | 2 |
| 71 | + | + | + | + | + | + | 6 |
| **Positive Results* n (%)** | **6 (14.3)** | **3 (7.1)** | **13 (31.0)** | **6 (14.3)** | **7 (16.7)** | **15 (35.7)** | **Total positive 17 (40.5%)** |

± indicates positive or negative amplification
*Percentages are presented as n/42 archived samples
Abbreviation: MLST, multi-locus sequence typing; PCR, polymerase chain reaction.

**Table S4.** Reference isolates from the *Leptospira* PubMLST database used in phylogenetic tree construction, including sequence types (STs) and allele profiles based on MLST scheme 3.

| ID | Isolate | Country | Year | Host | Species | Serovar | Serogroup | ST | Allele profile | | | | | |
|---|---|---|---|---|---|---|---|---|---|---|---|---|---|---|
| | | | | | | | | | ***adk*** | ***icdA*** | ***lipL32*** | ***lipL41*** | ***rrs*** | ***secY*** |
| 629 | IMRLEP-C18 | Malaysia | 2019 | Rat | *L. borgpetersenii* | NA | NA | 193 | 65 | 51 | 10 | 15 | 30 | 49 |
| 1514 | IMRLEP-K12 | Malaysia | 2019 | Rat | *L. borgpetersenii* | NA | NA | 194 | 65 | 76 | 10 | 15 | 30 | 49 |
| 884/ 1062 | Swart/ HAI0024 | Unknown | NA | NA | *L. interrogans* | Bataviae/ Canicola | NA | 149 | 5 | 1 | 2 | 3 | 1 | 5 |
| NA | NA | NA | NA | NA | *L. interrogans* | NA | NA | 154 | 5 | 2 | 2 | 50 | 2 | 13 |
| 143 | THAHKCHAN | India | NA | Human | *L. interrogans* | Other | Icterohaemorrhagiae | 3 | 1 | 1 | 2 | 2 | 1 | 43 |
| 93 | Butembo | Congo (DRC) | 1946 | Human | *L. kirschneri* | Butembo | Autumnalis | 100 | 17 | 24 | 11 | 16 | 7 | 5 |
| NA | NA | Indonesia | NA | NA | *L. kirschneri* | Cynopteri | Celledoni | 175 | 19 | 25 | 11 | 16 | 12 | 28 |
| 185 | 1348 U | Panama | NA | Rat | *L. noguchii* | Claytoni | Bataviae | 134 | 68 | 66 | 6 | 42 | 21 | 53 |
| 187 | Isolate 4 | Costa Rica | NA | Human | *L. noguchii* | Guaratuba | Pyrogenes | 135 | 68 | 68 | 6 | 44 | 7 | 4 |
| 192 | TRVL 112499 | Trinidad and Tobago | NA | Human | *L. santarosai* | Princestown | Pyrogenes | 110 | 25 | 30 | 15 | 25 | 10 | 32 |
| 106 | 735_U | Panama | 1966 | Rat | *L. santarosai* | Balboa | Bataviae | 114 | 30 | 35 | 19 | 23 | 10 | 37 |

Abbreviation: MLST, multi-locus sequence typing; NA, not available; ST, sequence type
